# Long-term healthcare resource use and cost associated with COVID-19 disease from a health system perspective. An equity-focused population-based cohort study

**DOI:** 10.64898/2026.01.16.26344255

**Authors:** A Ogunbameru, S Swayze, K Liu, S Mishra, B Sander

**Affiliations:** Institute of Health Policy, Management, and Evaluation, University of Toronto, Toronto, ON, Canada; Health Systems and Policy Research Collaborative Centre, University Health Network, Toronto, ON, Canada; ICES, Toronto, ON, Canada; Dalla Lana School of Public Health, University of Toronto, Toronto, ON, Canada; MAP Centre for Urban Health Solutions, Li Ka Shing Knowledge Institute, Unity Health Toronto, Toronto, ON, Canada; Department of Medicine and Institute of Medical Science, Temerty Faculty of Medicine, University of Toronto, Toronto, ON, Canada; Public Health Ontario, Toronto, ON, Canada

**Author notes:** Corresponding Author: Adeteju Ogunbameru, Health Systems and Policy Research Collaborative Centre, University Health Network, Toronto General Hospital, Eaton Building, 10th Floor, 200 Elizabeth Street, Toronto, ON M5G 2C4.

**Keywords:** COVID-19, Health system costs, Social determinants, Health disparities

## Abstract

SARS-CoV-2 strained Ontario’s health system, with social determinants of health (SDH) underexplored in cost analyses. We examined COVID-19 attributable healthcare resource use and costs from the Ontario health system perspective using health administrative data. We conducted a cohort study, matching 162,633 SARS-CoV-2-exposed individuals 1:1 to unexposed individuals. We calculated 10-day per-person mean attributable costs (2023 CAD) across care phases (pre-diagnosis, acute, post-acute, terminal), stratified by individual and area-level SDH. Among exposed individuals (mean age 40.4 years, 50.7% female), 6% were hospitalized, 1.3% admitted to critical care, and 2% died within 360 days. Mean (SD) person acute phase cost was $244 ($235-$253) and higher among males, recent immigrants, individuals living in low-income neighbourhoods and neighbourhoods with a higher proportion of crowded households. Extrapolating to the population level of 166,801 exposed individuals, the mean total survival-adjusted 360-day cost was $436 million. COVID-19 increased healthcare costs, disproportionately burdening marginalized communities.

## Introduction

From January 2020 to March 2024, 1,720,202 Ontario residents were reported to have been infected with SARS-CoV-2 (1). The COVID-19 pandemic significantly impacted the health of Ontarians, prompting the provincial government to invest over $10 billion in healthcare initiatives between 2020 and 2023, including funding for population and public health programs (e.g., testing and vaccination), personal protective equipment, additional hospital and long-term care beds, and support for frontline workers (2,3)

Preexisting social determinants of health (SDH) were amplified during the COVID-19 pandemic. In Ontario, residents of low-income and materially deprived neighbourhoods experienced higher rates of SARS-CoV-2 infection and mortality, while individuals who were recent immigrants or employed in essential jobs faced greater exposure and barriers to care (4). SDH have been shown to contribute to disparities in healthcare cost, e.g., an Ontario study found that individuals with stroke living in the most materially deprived neighbourhoods had significantly higher healthcare costs than those in the least deprived areas (11). underscoring how SDH contribute to disparities in health outcomes and resource use.

Stratifying healthcare costs by SDH provides insight into how healthcare resources are utilized across populations, which is critical for equitable health system planning.This approach is particularly important in the context of COVID-19, where marginalized populations have experienced higher infection rates, severe disease, and differential access to care, potentially leading to variations in healthcare utilization and associated costs (5).

Previous studies from England and Ontario have demonstrated that COVID-19-related healthcare costs vary by individual-level characteristics such as age, income level and co-morbidities (6–8). However, these studies lacked comprehensive stratification by SDH, limiting the understanding of cost disparities among marginalized groups. For example, Yang et al. (2023) reported substantial costs among people with severe COVID-19, particularly for older adults and those with comorbidities, but did not examine SDH (6). Tsui et al. (2022) compared initial healthcare costs among marginalized populations in British Columbia and Ontario, highlighting variation driven by differences in provincial healthcare systems, with acute care hospitalizations as the primary cost driver, yet long-term costs were not captured (7). Similarly, Sander et al. (2024) quantified short- and long-term healthcare costs in Ontario, identifying acute care hospitalizations as the main contributor, but provided limited data on SDH (9). These studies underscore the need for a deeper understanding of how SDH influence COVID-19 costs to inform healthcare delivery.

Our objective is to provide a comprehensive population-based analysis of COVID-19-attributable healthcare resource use and cost, stratified by SDH, from the Ontario health system perspective.

## Materials and methods

Our study methods followed the Reporting of Studies Conducted using Observational Routinely Collected Data guidelines (10).

### Study Design and Setting

We conducted a population-based, retrospective cohort study to estimate short- and long-term costs attributable to COVID-19 from the health system perspective (Ontario Ministry of Health) using an incidence-based costing approach. All data were sourced from administrative datasets housed at ICES, an independent, non-profit research institute. ICES links datasets using encoded identifiers, enabling health system evaluations without requiring patient consent under Ontario privacy laws (11). The ICES repository includes health service records of approximately 14 million Ontario residents (12).

### Population

The study included Ontario residents of all ages from January 1, 2016, to December 31, 2020, eligible for the Ontario Health Insurance Plan and with valid ICES identifiers. We excluded individuals with missing demographic data (e.g., sex, postal code, birth date), those aged >110 years, individuals with nosocomial COVID-19 infection, and long-term care residents, due to their distinct healthcare utilization patterns (13).

The exposed cohort comprised individuals with a positive polymerase chain reaction (PCR) test for SARS-CoV-2 in the COVID-19 Integrated Testing Data (C19INTGR) dataset between January 1 and December 31, 2020. The index date was defined as the date of the first positive PCR test.

The unexposed cohort consisted of Ontario residents identified from January 1, 2016, to December 31, 2018, with no positive COVID-19 test reported in C19INTGR dataset in 2020. We used a historical cohort to mitigate potential biases due to contamination (people who may have had COVID-19 but were not tested) and changes in health service utilization during 2020 (e.g., surgery cancellations) (14). Unexposed individuals were selected by randomly sampling 50% of the Ontario population from the Registered Persons Database (RPD) and were randomly assigned an index date based on the exposed cohort’s index date distribution.

Both cohorts were followed for one year post-index or until death.

Cohort selection is shown in Figure 1.

**Figure 1:**
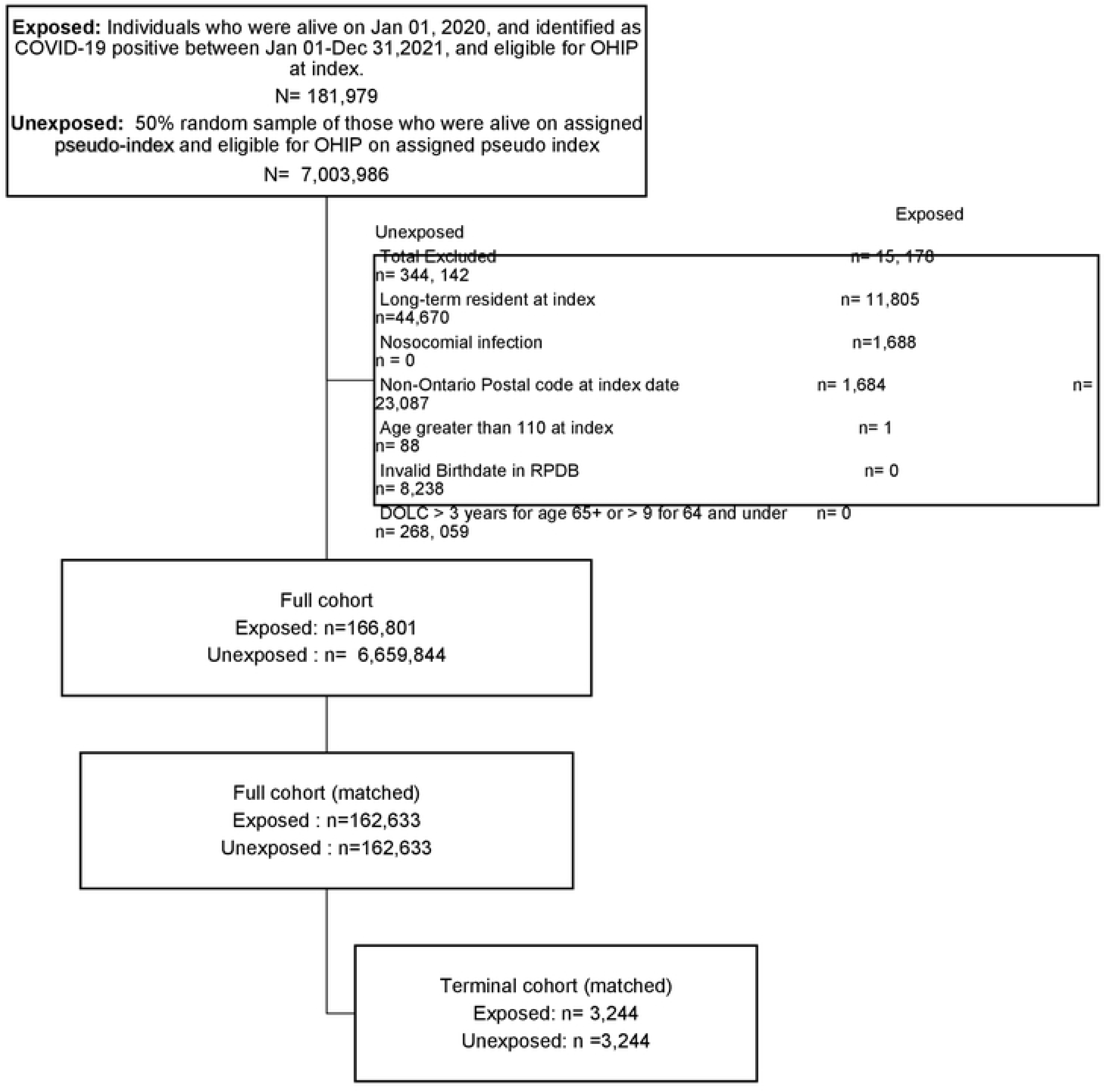
Cohort flowchart of exposed and unexposed populations

### Covariates and Data Sources

We linked demographic, clinical, and social determinants of health (SDH) data from C19INTGR and other ICES datasets using unique encoded identifiers (15). Baseline characteristics included demographic and clinical factors (age, sex, comorbidity, rurality) and social determinants of health (SDH), defined at the individual level (e.g., immigration status) and area level (e.g., neighbourhood income, household crowding, proportion of essential workers).

Comorbidity was measured as the Resource Utilization Band based on the Johns Hopkins Aggregated Diagnosis Groups two years prior to index date (16). Age, sex, and rurality were sourced from RPD. Immigration status was obtained from the Immigration, Refugees, and Citizenship Canada Permanent Resident Database in ICES.

For area-level SDH, we considered variables previously shown to be associated with increased odds of SARS-CoV-2 infection (17) or higher healthcare costs in Ontario (18,19). We included neighbourhood income quintile, % visible minority living in an area, household crowding index (number of persons per room), and % working in essential services in an area. Area-level SDH data were sourced from the 2016 Canadian Census at the Dissemination Area (DA) level, a small, relatively stable geographic unit with an average population of 400–700 persons. DA-level data were assigned to individuals based on residential postal codes (20).

All baseline characteristics were assessed at the index date. Details and definitions for each variable are included in Appendix Table A1.

Healthcare utilization and cost data were sourced from multiple datasets (15) including the Canadian Institute for Health Information’s Discharge Abstract Database (acute inpatient hospitalization), Ontario Mental Health Reporting System (mental health), Continuing Care Reporting System (complex continuing care), National Rehabilitation System (inpatient rehabilitation), National Ambulatory Care Reporting System (same day surgeries), Ontario Health Insurance Plan (physician services), Ontario Drug Benefit (prescription drugs), and Office of the Registrar General Death registry.

Data was assessed between March 1, 2022 and December 31, 2024. Authors had access to information that could identify individual participants during or after data collection.

### Outcomes

Our primary outcome were COVID-19-attributable costs by phase of care, standardized to mean 10-day per-person costs in 2023 Canadian dollars. Addtional outcomes included all-cause mortality and resource use (e.g., hospitalizations, intensive care unit (ICU) admissions) among the exposed cohort.

### Matched Cohort

We matched each exposed individual to one unexposed individual using hard- and propensity score matching.

We calculated propensity scores using greedy matching with a caliper width of 0.2 standard deviations (SD) (21). Covariates included in the propensity score model were individual-level (rurality, immigration status) and area-level variables (marginalization indices, housing, occupation, and visible minority status) measured at index date. Hard matching was performed on sex, same age, index date (±60 days), and logit of the propensity score. Balance between groups was assessed using weighted standardized differences with <0.1 indicating negligible imbalance (21). Variables included in propensity score model are described in Appendix Table A1.

For stratified analyses, the variable of interest was excluded from the propensity score model and included in the hard match alongside sex, age, index date (± 60 days) and logit of the propensity score.

To estimate terminal phase costs, we matched deceased exposed individuals to unexposed individuals of the same age or older to better account for predictors of cost prior to death. Other variables included in this match were sex, logit of the propensity score with a caliper width of 0.2 SD (calculated at index), and death date ±30 days. Terminal phase costs were stratified by early (≤60 days post-index) and late deaths (>60 days post-index).

### Analysis

Healthcare resource use (hospitalizations, ICU admissions, mortality) is reported for exposed individuals prior to matching. Continuous variables are reported as mean (SD) and median (IQR), categorical variables as frequencies and percentages.

Post-matching, costs, standardized to mean 10-day per-person cost, were calculated and plotted over 360 days, pre- and post-index date. Four care phases were defined using joinpoint analysis, guided by clinical expertise (SM and BS) and the WHO’s post-COVID-19 condition framework (22): pre-diagnosis (20 days pre-index), acute (80 days post-index), post-acute (days 81–360), and terminal (60 days prior to death date) (Appendix Figure A1).

Phase-specific attributable costs were calculated as the mean cost differences (with 95% confidence intervals (95% CI)) between matched exposed and unexposed individuals using a generalized estimating equation model.

Costs were stratified by individual- and area-level SDH (age, sex, immigration status, household crowding index, %visible minority living in an area, and % essential workers living in an area).

Phase-specific costs were combined with crude survival data to estimate survival-adjusted mean (minimum and maximum) per-person costs at 30, 90, 180, and 360 days (23), and stratified by sex.

All analyses were conducted using SAS Enterprise Guide 8.3 (SAS Institute, Inc.).

## Ethics approval

This study was approved by the University Health Network Research Ethics Board. ICES is a prescribed entity under Ontario’s Personal Health Information Protection Act (PHIPA). Section 45 of the PHIPA authorizes the ICES to collect personal health information, without consent, for the purpose of analysis or compiling statistical information with respect to the management of, evaluation or monitoring of the allocation of resources to, or planning for all or part of the health system. Projects that use data collected by ICES under section 45 of PHIPA and use no other data are exempt from the REB review. The use of the data in this project is authorized under Section 45 and approved by the ICES Privacy and Legal Office.

## Results

A cohort of 166,801 individuals with confirmed COVID-19 met the study inclusion criteria (Figure 1).

Their mean (SD) age was 40.38 (19.70) years, 50.6% were female. Most (96%) lived in urban areas, 21.4% were high healthcare resource users, 46.2% resided in low-income neighbourhoods (quintiles 1& 2), 8.3% were recent immigrants (migrated to Canada within five years), and 42.1% lived in crowded dwellings (2 or more persons per room). The unexposed cohort of 6,659,844 individuals had a mean (SD) age of 40.5 (22.7) years, 50.6% were female. Of these, 89.3% lived in urban areas, 17.1% were high healthcare resource users, 39.1% resided in low-income neighbourhoods (quintiles 1&2), 3.4% were recent immigrants, and 22.6% lived in crowded dwellings (2 or more persons per room).

Baseline characteristics are presented in Table 1.

**Table 1:** Baseline characteristics of exposed and unexposed groups before and after matching.

| Matching Variables | Before |  |  | After matching |  |  |
| --- | --- | --- | --- | --- | --- | --- |
|  | Exposed cohort<br>N=166,801 | Unexposed cohort<br>N=6,659,844 | Weighted standardized difference | Exposed cohort<br>N=162,633 | Unexposed cohort<br>N=162,633 | Weighted standardized difference |
| **Age at index** |  |  |  |  |  |  |
| Mean ± SD | 40.38 ± 19.70 | 40.49 ± 22.75 | 0.01 | 40.44 ± 19.77 | 40.39 ± 19.74 | 0 |
| Median (IQR) | 39 (25-55) | 41 (22-58) | 0.01 | 39 (25-55) | 39 (25-55) | 0 |
| **Age category at Index n (%)** |  |  |  |  |  |  |
| 0 to less than 4 years | 3,355 (2.0) | 350,617 (5.3) | 0.17 | 3,325 (2.0) | 3,325 (2.0) | 0 |
| 5years to 10 years | 5,640 (3.3) | 448,166 (6.7) | 0.15 | 5,586 (3.4) | 5,586 (3.4) | 0 |
| 11 years to 15 years | 6,497 (3.9) | 364,842 (5.5) | 0.08 | 6,412 (3.9) | 6,412 (3.9) | 0 |
| 16 years to 20 years | 10,792 (6.4) | 372,118 (5.6) | 0.04 | 10,532 (6.5) | 10,532 (6.5) | 0 |
| 21 years to 25 years | 16,508 (9.8) | 422,873 (6.3) | 0.13 | 15,761 (9.7) | 15,761 (9.7) | 0 |
| 26years to 30 years | 17,128 (10.2) | 455,097 (6.8) | 0.12 | 16,387 (10.1) | 16,387 (10.1) | 0 |
| 31 years to 35 years | 14,503 (8.6) | 455,917 (6.8) | 0.07 | 14,011 (8.6) | 14,011 (8.6) | 0 |
| 36years to 40 years | 13,311 (7.9) | 436,193 (6.5) | 0.05 | 12,896 (7.9) | 12,896 (7.9) | 0 |
| 41years to 45 years | 12,777 (7.6) | 441,210 (6.6) | 0.04 | 12,392 (7.6) | 12,392 (7.6) | 0 |
| 46 years to 50 years | 13,532<br>(8.0) | 468,336<br>(7.0) | 0.04 | 13,133<br>(8.1) | 13,133<br>(8.1) | 0 |
| 51years to 55 years | 13,875<br>(8.2) | 518,510<br>(7.8) | 0.02 | 13,479<br>(8.3) | 13,479<br>(8.3) | 0 |
| 56years to 60 years | 12,865<br>(7.6) | 486,774<br>(7.3) | 0.01 | 12,480<br>(7.7) | 12,480<br>(7.7) | 0 |
| 61years to 65 years | 9,566 (5.7) | 417,436<br>(6.3) | 0.03 | 9,264 (5.7) | 9,264 (5.7) | 0 |
| 66years to 70 years | 5,772 (3.4) | 353,134<br>(5.3) | 0.09 | 5,582 (3.4) | 5,582 (3.4) | 0 |
| 71years to 75years | 4,124 (2.4) | 256,530<br>(3.9) | 0.08 | 3,922 (2.4) | 3,922 (2.4) | 0 |
| 76years to 80 years | 2,829 (1.7) | 181,928<br>(2.7) | 0.08 | 2,631 (1.6) | 2,631 (1.6) | 0 |
| 81years + | 5,415 (3.2) | 230,163<br>(3.5) | 0.03 | 4,840 (3.0) | 4,840 (3.0) | 0 |
| **Rurality n (%)** |  |  |  |  |  |  |
| No | 160,181<br>(96.0) | 5,946,912<br>(89.3) | 0.26 | 156,039<br>(95.9) | 155,891<br>(95.9) | 0 |
| Yes | 5,695 (3.4) | 696,606<br>(10.5) | 0.28 | 5,678 (3.5) | 5,830 (3.6) | 0.01 |
| Missing | 925 (0.6) | 16,326 (0.2) | 0.05 | 916 (0.6) | 912 (0.6) | 0 |
| **Sex n (%)** |  |  |  |  |  |  |
| Female | 84,449<br>(50.6) | 3,371,522 (50.6) |  | 82,418<br>(50.7) | 82,418<br>(50.7) | 0 |
| Male | 82,352<br>(49.4) | 3,288,322 (49.4) |  | 80,215<br>(49.3) | 80,215<br>(49.3) | 0 |
| **Senior at Index n (%)** |  |  |  |  |  |  |
| Yes | 17,201<br>(10.3) | 1,021,755<br>(15.3) | 0.15 | 16,975<br>(10.4) | 16,975<br>(10.4) | 0 |
| **Average total household income n (%) \*** |  |  |  |  |  |  |
| 1st quintile (lowest) | 34,203<br>(20.5) | 1,233,408<br>(18.5) | 0.05 | 33,574<br>(20.6) | 33,734<br>(20.7) | 0 |
| 2nd quintile | 28,885<br>(17.3) | 1,243,533<br>(18.7) | 0.04 | 28,203<br>(17.3) | 28,456<br>(17.5) | 0 |
| 3rd quintile | 29,609<br>(17.8) | 1,235,749<br>(18.6) | 0.02 | 28,743<br>(17.7) | 28,551<br>(17.6) | 0 |
| 4th quintile | 42,021<br>(25.2) | 1,507,082<br>(22.6) | 0.06 | 40,629<br>(25.0) | 40,075<br>(24.6) | 0.01 |
| 5th quintile | 30,937<br>(18.5) | 1,409,153<br>(21.2) | 0.07 | 30,347<br>(18.7) | 30,705<br>(18.9) | 0.01 |
| Missing | 1,146 (0.7) | 30,919 (0.4) | 0.07 | 1,137 (0.7) | 1,112 (0.7) | 0 |
| **Material deprivation quintile n (%)** |  |  |  |  |  |  |
| 1st quintile (highest) | 28,181<br>(16.9) | 1,512,067<br>(22.7) | 0.15 | 27,898<br>(17.2) | 27,740<br>(17.1) | 0 |
| 2nd quintile | 29,011<br>(17.4) | 1,390,700<br>(20.9) | 0.09 | 28,500<br>(17.5) | 28,153<br>(17.3) | 0.01 |
| 3rd quintile | 32,677<br>(19.6) | 1,258,647<br>(18.9) | 0.02 | 31,728<br>(19.5) | 31,548<br>(19.4) | 0 |
| 4th quintile | 34,573<br>(20.7) | 1,204,520<br>(18.1) | 0.07 | 33,398<br>(20.5) | 33,513<br>(20.6) | 0 |
| 5th quintile | 41,284<br>(24.8) | 1,230,371<br>(18.5) | 0.15 | 40,278<br>(24.8) | 40,080<br>(24.6) | 0 |
| Missing | 1,075 (0.6) | 63,539 (1.0) | 0.03 | 831 (0.5) | 1,599 (1.0) | 0.05 |
| **% dwellings in need of major repair n (%) \*** |  |  |  |  |  |  |
| 1st quartile (highest) | 85,024<br>(51.0) | 3,114,465<br>(46.8) | 0.08 | 82,574<br>(50.8) | 82,116<br>(50.5) | 0.01 |
| 2nd quartile | 26,608<br>(16.0) | 1,183,620<br>(17.8) | 0.05 | 25,969<br>(16.0) | 26,204<br>(16.1) | 0 |
| 3rd quartile | 24,977<br>(15.0) | 1,116,629<br>(16.8) | 0.05 | 24,422<br>(15.0) | 24,553<br>(15.1) | 0 |
| 4th quartile | 26,709<br>(16.0) | 1,097,647<br>(16.5) | 0.01 | 26,253<br>(16.1) | 26,448<br>(16.3) | 0 |
| Missing | 3,483 (2.1) | 147,483<br>(2.2) | 0.08 | 3,415 (2.1) | 3,313 (2.7) | 0.01 |
| **% essential workers n (%) \*** |  |  |  |  |  |  |
| 1st quintile (lowest) | 26,206<br>(15.7) | 1,412,124<br>(21.2) | 0.14 | 26,009<br>(16.0) | 25,768<br>(15.8) | 0 |
| 2nd quintile | 34,294<br>(20.6) | 1,473,344<br>(22.1) | 0.04 | 33,675<br>(20.7) | 33,686<br>(20.7) | 0 |
| 3rd quintile | 32,255<br>(19.3) | 1,312,734<br>(19.7) | 0.01 | 31,385<br>(19.3) | 31,818<br>(19.6) | 0.01 |
| 4th quintile | 35,589<br>(21.3) | 1,268,151<br>(19.0) | 0.06 | 34,393<br>(21.1) | 34,250<br>(21.1) | 0 |
| 5th quintile | 37,311<br>(22.4) | 1,162,463<br>(17.5) | 0.12 | 36,034<br>(22.2) | 35,999<br>(22.1) | 0 |
| Missing | 1,146 (0.7) | 31,028 (0.5) | 0.03 | 1,137 (0.7) | 1,112 (0.7) | 0 |
| **Household Crowding Index n (%)** |  |  |  |  |  |  |
| One person or fewer per room | 93,752<br>(56.2) | 5,059,871<br>(76.0) | 0.43 | 92,482<br>(56.9) | 92,568<br>(56.9) | 0 |
| Two or more persons per room | 70,306<br>(42.1) | 1,507,733<br>(22.6) | 0.43 | 67,462<br>(41.5) | 67,461<br>(41.5) | 0 |
| Missing | 2,743 (1.7) | 92,240 (1.3) | 0.05 | 2,689 (1.7) | 2,604 (1.7) | 0.01 |
| **Recent immigrant** |  |  |  |  |  |  |
| No | 6,435,781<br>(96.6) | 153,023<br>(91.7) | 0.21 | 149,675<br>(92.0) | 149,675<br>(92.0) | 0 |
| Yes | 224,063<br>(3.4) | 13,778 (8.3) | 0.21 | 13,014<br>(8.0) | 13,014<br>(8.0) | 0 |
| **% lone parent families n (%) \*** |  |  |  |  |  |  |
| 1st quintile (lowest) | 22,552<br>(13.5) | 1,246,261<br>(18.7) | 0.14 | 22,145<br>(13.6) | 21,646<br>(13.3) | 0.01 |
| 2nd quintile | 35,004<br>(21.0) | 1,442,293<br>(21.7) | 0.02 | 33,939<br>(20.9) | 33,654<br>(20.7) | 0 |
| 3rd quintile | 34,125<br>(20.5) | 1,365,710<br>(20.5) | 0 | 33,155<br>(20.4) | 33,234<br>(20.4) | 0 |
| 4th quintile | 32,536<br>(19.5) | 1,261,037<br>(18.9) | 0.01 | 31,656<br>(19.5) | 32,299<br>(19.9) | 0.01 |
| 5th quintile | 41,356<br>(24.8) | 1,310,563<br>(19.7) | 0.12 | 40,519<br>(24.9) | 40,581<br>(25.0) | 0 |
| Missing | 1,228 (0.8) | 33,980 (0.5) | 0.02 | 1,219 (0.8) | 1,219 (0.8) | 0 |
| **Neighbourhood income quintile n (%) \*** |  |  |  |  |  |  |
| 1st quintile (lowest) | 40,878<br>(24.5) | 1,298,211<br>(19.5) | 0.12 | 39,944<br>(24.6) | 40,118<br>(24.7) | 0 |
| 2nd quintile | 36,212<br>(21.7) | 1,303,219<br>(19.6) | 0.05 | 35,014<br>(21.5) | 35,211<br>(21.7) | 0 |
| 3rd quintile | 36,205<br>(21.7) | 1,331,629<br>(20.0) | 0.04 | 34,980<br>(21.5) | 34,899<br>(21.5) | 0 |
| 4th quintile | 29,252<br>(17.5) | 1,342,461<br>(20.2) | 0.07 | 28,689<br>(17.6) | 28,424<br>(17.5) | 0 |
| 5th quintile | 23,291<br>(14.0) | 1,365,554<br>(20.5) | 0.17 | 23,053<br>(14.2) | 23,026<br>(14.2) | 0 |
| Missing | 963 (0.6) | 18,770 (0.3) | 0.05 | 953 (0.6) | 955 (0.6) | 0 |
| **% living in suitable housing n (%) \*** |  |  |  |  |  |  |
| 1st quintile (lowest) | 71,095<br>(42.6) | 1,537,412<br>(23.1) | 0.43 | 68,420<br>(42.1) | 68,562<br>(42.2) | 0 |
| 2nd quintile | 33,741<br>(20.2) | 1,321,899<br>(19.8) | 0.01 | 32,952<br>(20.3) | 32,714<br>(20.1) | 0 |
| 3rd quintile | 24,704<br>(14.8) | 1,269,778<br>(19.1) | 0.11 | 24,335<br>(15.0) | 24,121<br>(14.8) | 0 |
| 4th quintile | 15,817<br>(9.5) | 1,092,549<br>(16.4) | 0.21 | 15,692<br>(9.6) | 15,914<br>(9.8) | 0 |
| 5th quintile | 18,545<br>(11.1) | 1,316,216<br>(19.8) | 0.24 | 18,372<br>(11.3) | 18,544<br>(11.4) | 0 |
| Missing | 2,899 (1.8) | 121,990<br>(2.0) | 0.03 | 2,862 (1.8) | 2,778 (1.7) | 0 |
| **% self-identify as visible minority n (%) \*** |  |  |  |  |  |  |
| 1st quintile (lowest) | 9,367 (5.6) | 1,060,963<br>(15.9) | 0.34 | 9,338 (5.7) | 9,415 (5.8) | 0 |
| 2nd quintile | 12,878<br>(7.7) | 1,121,844<br>(16.8) | 0.28 | 12,840<br>(7.9) | 12,900<br>(7.9) | 0 |
| 3rd quintile | 20,254<br>(12.1) | 1,195,814<br>(18.0) | 0.16 | 20,095<br>(12.4) | 20,233<br>(12.4) | 0 |
| 4th quintile | 37,013<br>(22.2) | 1,433,418<br>(21.5) | 0.02 | 36,404<br>(22.4) | 36,068<br>(22.2) | 0 |
| 5th quintile | 86,143<br>(51.6) | 1,816,886<br>(27.3) | 0.51 | 82,819<br>(50.9) | 82,905<br>(51.0) | 0 |
| Missing | 1,146 (0.7) | 30,919 (0.4) | 0.02 | 1,137 (0.7) | 1,112 (0.7) | 0 |
| **Resource Utilization band n (%)** |  |  |  |  |  |  |
| Non-users | 12,680<br>(7.6) | 801,768<br>(12.0) | 0.15 | 12,475<br>(7.7) | 12,366<br>(7.6) | 0 |
| Healthy Users | 8,384 (5.0) | 413,821<br>(6.2) | 0.05 | 8,218 (5.1) | 7,765 (4.8) | 0.01 |
| Low resource utilization | 29,166<br>(17.5) | 1,330,796<br>(20.0) | 0.06 | 28,581<br>(17.6) | 27,940<br>(17.2) | 0.01 |
| Moderate resource utilization | 81,794<br>(49.0) | 2,977,328<br>(44.7) | 0.09 | 79,922<br>(49.1) | 81,094<br>(49.9) | 0.01 |
| High resource utilization | 25,180<br>(15.1) | 825,920<br>(12.4) | 0.08 | 24,257<br>(14.9) | 24,369<br>(15.0) | 0 |
| Very high resource utilization | 9,597 (5.8) | 310,211<br>(4.7) | 0.05 | 9,180 (5.6) | 9,099 (5.6) | 0 |
| **All-cause mortality n (%)** |  |  |  |  |  |  |
| Yes | 3,375 (2.0) | 214,800<br>(3.2) | 0.08 | 3,304 (2.0) | 4,438 (2.7) | 0.05 |
| **COVID-19 related hospitalization n (%)** |  |  |  |  |  |  |
| Yes | 10,000<br>(6.0) | 0 (0.0) |  | NR | NR |  |
| **COVID-19-related ICU admission n (%)** |  |  |  |  |  |  |
| Yes | 2,131 (1.3) | 0 (0.0) |  | NR | NR |  |

IQR: interquartile range; SD: standard deviation; NA: not required; \* Dissemination area-level variables from Stats Canada, indicating the proportion of adults/households with the variable of interest (calculated in quartiles/quintiles) (see a more detailed description of each variable in Appendix Table A

### Healthcare Utilization

Within the 360-day follow-up, 5.9% of individuals with confirmed COVID-19 were hospitalized, 1.3% required ICU admission, and 2% died (all-cause mortality).

Among individuals admitted to ICU, 49.9% were aged >65 years, 65.4% were male, 89.6% were within the moderate to very high healthcare resource utilization band, and 57.2% resided in low-income neighbourhoods (quintiles 1& 2).

Among individuals who died, 48.8% were aged >80 years, 55.1% were male, 51.1% resided in low-income neighbourhoods (quintiles 1&2), and 63.3% died within 60 days of confirmed COVID-19 (early deaths).

The median (IQR) hospital length of stay for late deaths was 239 days (55, 627).

Of the 166,801 exposed individuals who met the study inclusion criteria, 97.5% were matched 1:1 to unexposed individuals with standardized differences < 0.1, indicating good balance (Table 1).

### Healthcare Costs

#### Full cohort Analysis

The mean (95% CI) total COVID-19 attributable 10-day per-person phase-specific cost for the pre-diagnosis, acute, post-acute and terminal care phase (combined early and late deaths) were $11 ($6-$15), $244 ($235-$253), $20 ($17-$24), and $4,753 ($4,419-$5,088), respectively (Table 2)

**Table 2:**
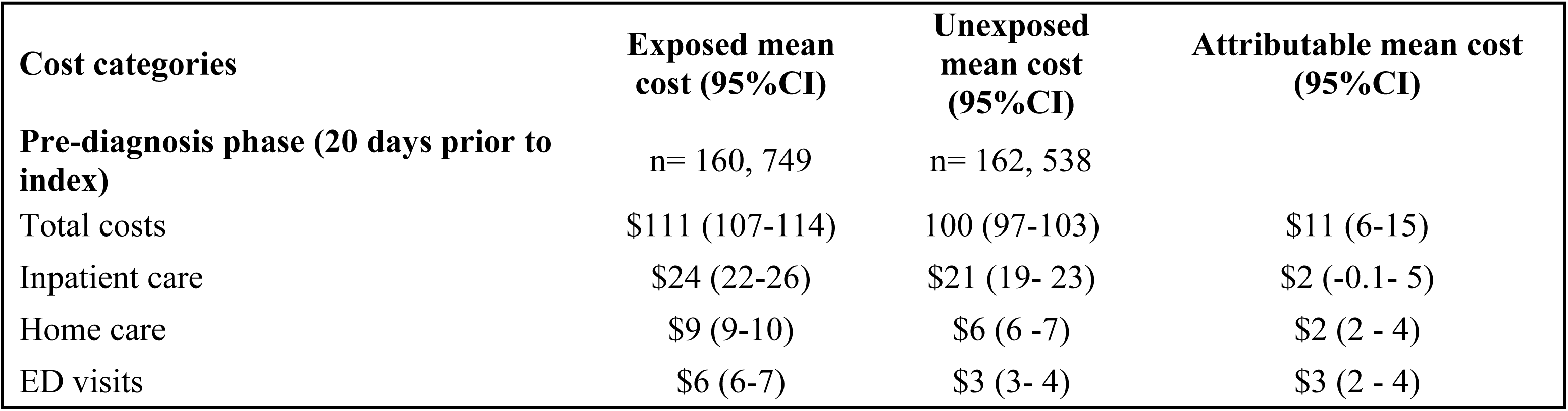
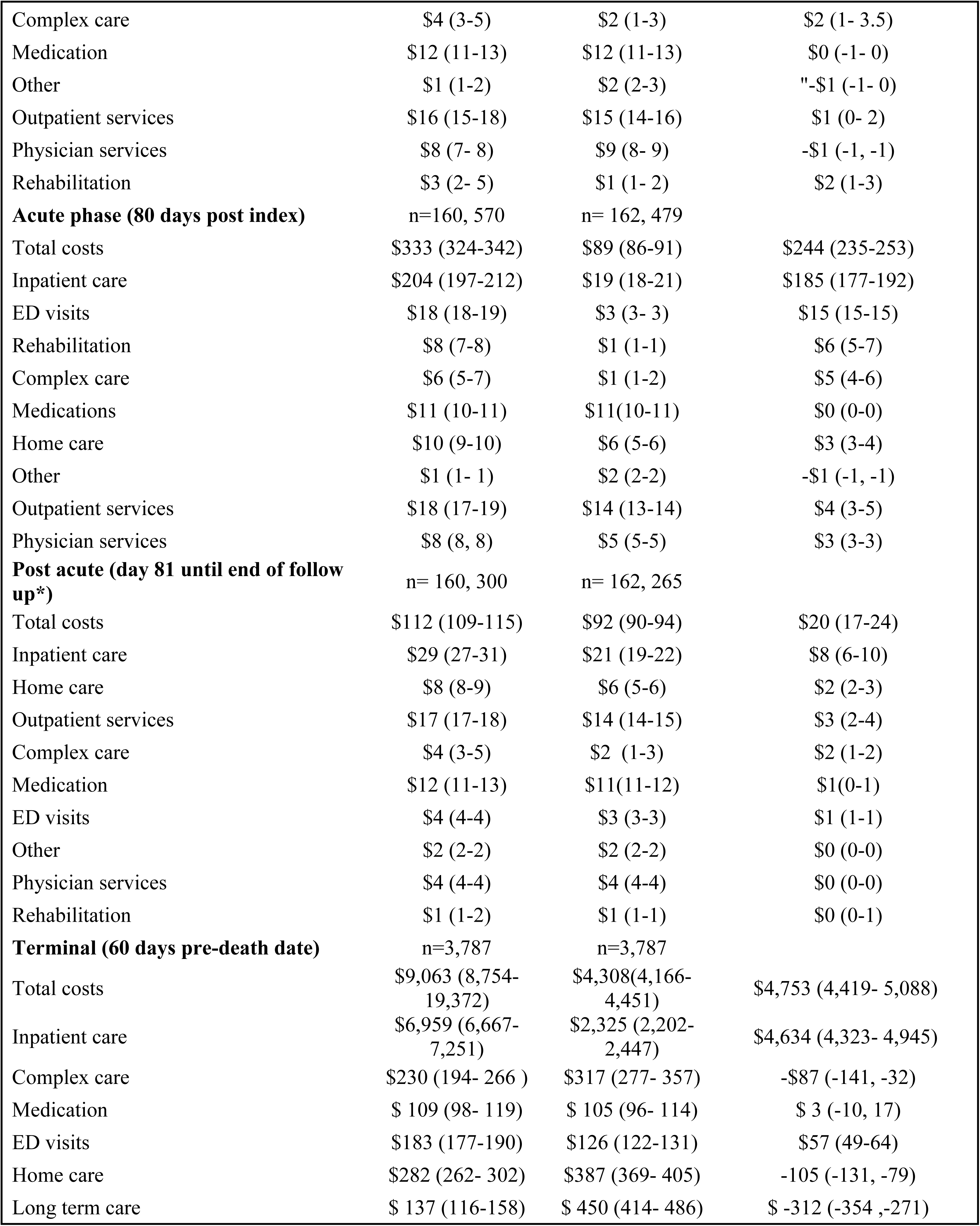
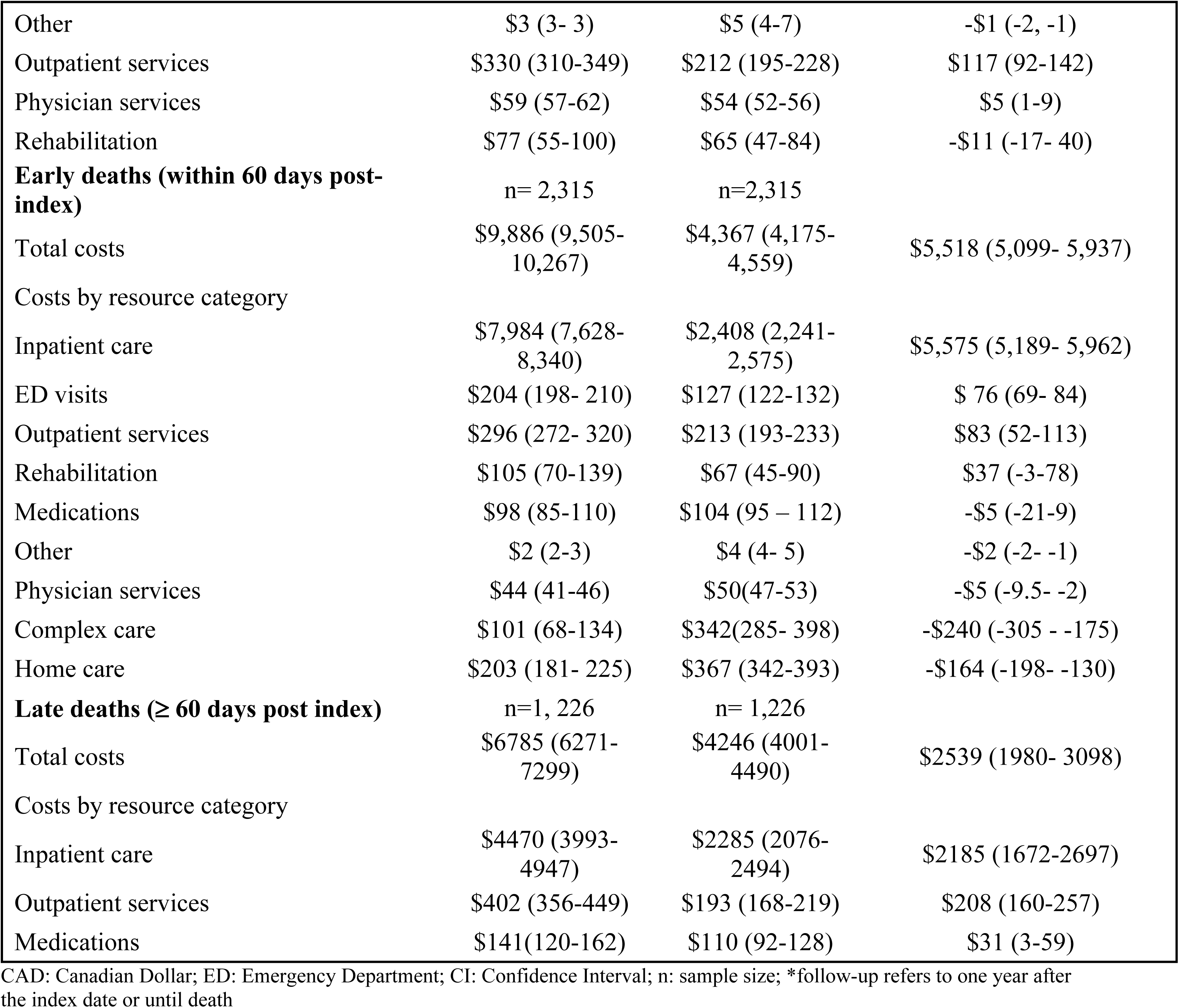
COVID-19-attributable healthcare costs (2023 CAD) stratified by phase of care.

Cost components per phase are shown in Figure 2.

**Figure 2:**
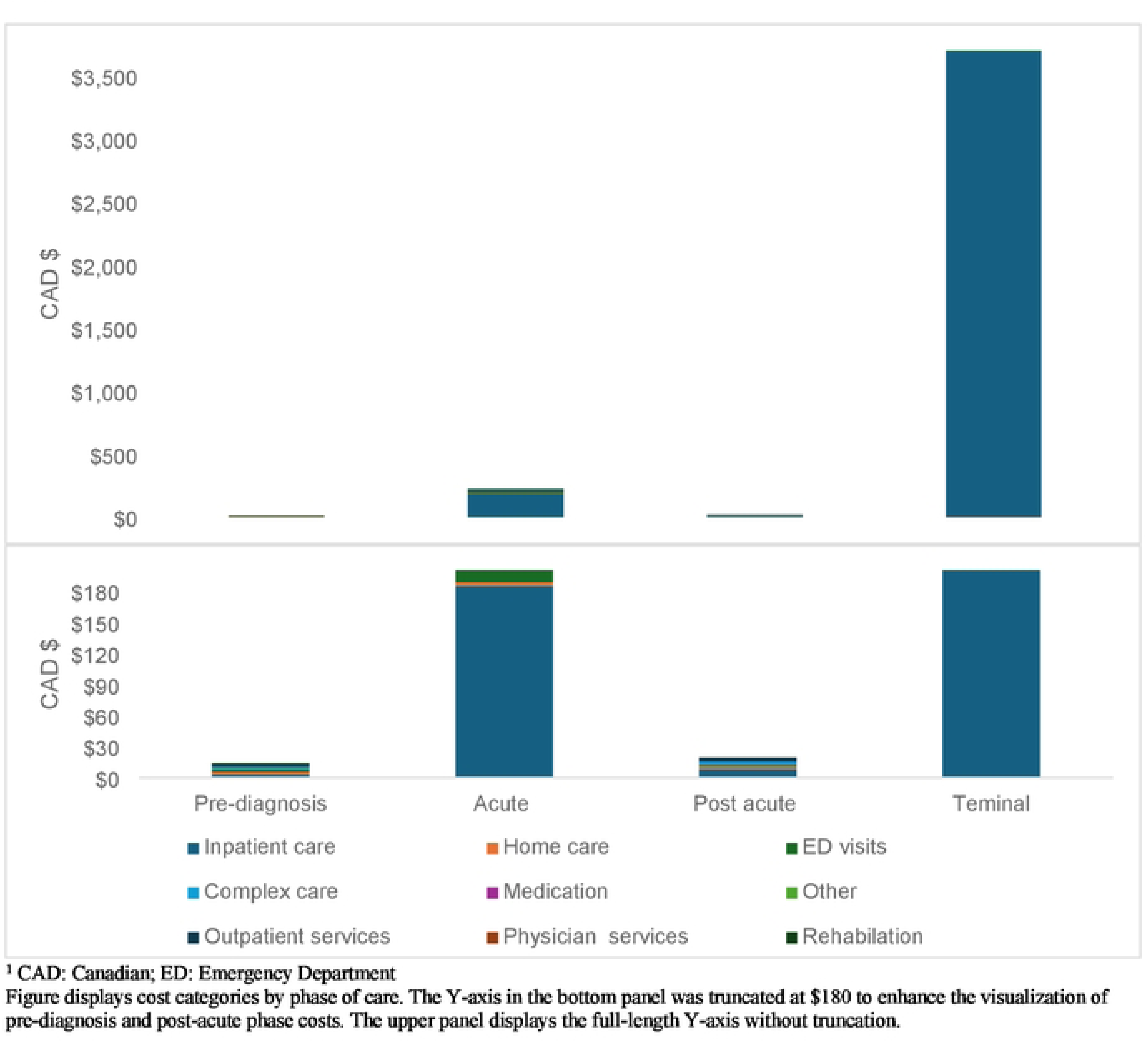
Total Attributable COVID-19 cost (in 2023 CAD) per healthcare service components per phase of care

In the pre-diagnosis care phase, hospitalizations, home care services and emergency department visits were the main contributors to the mean phase cost, each accounting for approximately 27% of the mean total phase cost at $3 ($0-$6), $3 ($2-$4), and $3 ($2-$3), respectively.

In the acute care phase, hospitalizations were the largest cost component at $185 ($177-$192), accounting for 76% of the mean total cost. Emergency department visits contributed 6% to the mean total phase cost, and rehabilitation contributed 2%.

In the post-acute phase, hospitalizations contributed 40% to the mean phase costs at $8 ($6-$11), followed by outpatient visits (15%) at $3 ($2-$4), and home care services (10%) at $2 ($2-$3).

In the terminal phase, mean (95% CI) attributable costs were higher for early deaths ($5,518 [$5,099–$5,937]) than for late deaths ($3,549 [$3,001–$4,097]). For early deaths, hospitalizations accounted for 100% of the mean total cost ($5,575 [$5,189–$5,962]). For late deaths, hospitalizations accounted for 88.9% of the mean total cost at $3,152 ($2,642–$3,662), with home care costs ($6 [−$41 to $54]) and complex care costs ($14 [−$87 to $117]) being other major cost categories. (Table 2).

#### Stratified Analysis

Males had higher costs in the acute ($293 [278-$308]) and terminal phases ($5,690 [$5,206–$6,174]) compared to females (acute: $197 [$186- $207], terminal: ($3,569 [$3,129–$4,008]) (Table 3).

**Table 3:** Phase-specific COVID-19 per-person costs (2023 CAD) stratified by sex.

| Sex | Exposed (n) | Exposed mean cost (95% CI) | Unexposed (n) | Unexposed mean cost (95% CI) | Attributable mean cost (95% CI) |
| --- | --- | --- | --- | --- | --- |
| **Pre-diagnosis phase (20 days prior to index date)** |  |  |  |  |  |
| Female | 81,647 | \$122 (117-127) | 82,372 | \$108 (103-112) | \$14.6 (7-21) |
| Male | 79,102 | \$99 (94-104) | 80,166 | \$92 (87-97) | \$ 7 (1-14) |
| **Acute phase (80 days post index)** |  |  |  |  |  |
| Female | 81,582 | \$293(283-304) | 82,344 | \$96 (93-100) | \$197 (186-207) |
| Male | 78,988 | \$374 (359-389) | 80,135 | \$80 (77- 84) | \$293 (278-308) |
| **Post acute (day 81 until end of follow up\*)** |  |  |  |  |  |
| Female | 81,455 | \$120 (116-124) | 82,249 | \$98 (95-100) | \$23 (18-27) |
| Male | 78,845 | \$104 (100-109) | 80,016 | \$86 (83-89) | \$18 (13-23) |
| **Terminal phase (60 days pre-death date)** |  |  |  |  |  |
| Female | 1,673 | \$7,602 (7,191-8,012) | 1,680 | \$ 4,033 (3,850-4,216) | \$ 3,569 (3,129-4,008) |
| Male | 2,114 | \$ 10,219 (9,776-10,661) | 2,112 | \$4,528 (4,318-4,738) | \$5,690 (5,206- 6,174) |
| Female | 1,673 | \$7,602 (7,191-8,012) | 1,680 | \$ 4,033 (3,850-4,216) | \$ 3,569 (3,129-4,008) |
| Male | 2,114 | \$ 10,219 (9,776-10,661) | 2,112 | \$4,528 (4,318-4,738) | \$5,690 (5,206- 6,174) |
| Female | 1,673 | \$7,602 (7,191-8,012) | 1,680 | \$ 4,033 (3,850-4,216) | \$ 3,569 (3,129-4,008) |

Individuals aged ≥70 years had higher costs compared to other age groups in the pre-diagnosis, acute care, post-acute care and terminal phases (Table 4).

**Table 4:** Phase-specific COVID-19 per-person costs (2023 CAD) stratified by age.

| Age group | Exposed (n) | Exposed mean cost (95% CI) | Unexposed (n) | Unexposed mean cost (95% CI) | Attributable cost mean (95% CI) |
| --- | --- | --- | --- | --- | --- |
| **Pre-diagnosis phase (20 days prior to index)** |  |  |  |  |  |
| Less than 2 | 1,141 | \$98 (65-132) | 1,341 | \$121 (74-168) | -\$23(-80-34) |
| 2-4 | 2,183 | \$28(17- 38) | 1,984 | \$39 (28-51) | -\$11 (-26- 4) |
| 5-11 | 6,734 | \$22(18- 26) | 6,916 | \$27 (22-33) | -\$5 (-12- 1) |
| 12-17 | 8,632 | \$34 (27- 40) | 9,082 | \$35 (29-40) | -\$1 (-8- 7) |
| 18-29 | 36,153 | \$43 (39-47) | 35,373 | \$49 (45-53) | -\$6 (-12- -1) |
| 30-49 | 52,803 | \$65 (61-69) | 53,013 | \$68 (64-72) | -\$3 (-8-2) |
| 50- 69 | 42,190 | \$137 (128-145) | 42,467 | \$131 (123-138) | \$5 (-5-16) |
| 70 and above | 10,913 | \$591 (555-627) | 12,362 | \$371 (348-393) | \$220 (178-263) |
| **Acute phase (80 days post index)** |  |  |  |  |  |
| Less than 2 | 1,141 | \$189 (151-228) | 1,341 | \$50 (35- 65) | \$139 (97-180) |
| 2-4 | 2,183 | \$49 (37- 61) | 1,984 | \$33 (26-40) | \$16 (2-30) |
| 5-11 | 6,734 | \$43 (32-54) | 6,916 | \$25 (21-30) | \$17 (6-29) |
| 12-17 | 8,632 | \$47 (41-54) | 9,082 | \$34 (30-38) | \$13 (5-21) |
| 18-29 | 36,152 | \$71 (66-76) | 35,371 | \$42 (39-45) | \$29 (23- 35) |
| 30-49 | 52,799 | \$169 (159-179) | 53,008 | \$59 (56-61) | \$110 (99-120) |
| 50- 69 | 42,138 | \$531 (506-555) | 42,451 | \$115 (110-120) | \$415 (390-440) |
| 70 and above | 10,791 | \$1723 (1652-1795) | 12,326 | \$349 (330-367) | \$1375 (1302-1448) |
| **Post acute (day 81 until end of follow up\*)** |  |  |  |  |  |
| Less than 2 | 1,141 | \$51(36- 65) | 1,341 | \$33 (26- 41) | \$17 (1-33) |
| 2-4 | 2,183 | \$39 (19- 58) | 1,984 | \$26 (20-33) | \$12 (-8, 32) |
| 5-11 | 6,734 | \$21 (17- 24) | 6,916 | \$27(21- 32) | -\$5 (-12, 1) |
| 12-17 | 8,632 | \$33 (28-38) | 9,082 | \$31 (27- 34) | \$2 (-3, 8) |
| 18-29 | 36,147 | \$44 (41-47) | 35,366 | \$44 (42-47) | -\$1 (-4, 3) |
| 30-49 | 52,788 | \$68 (65-72) | 52,989 | \$59 (57-61) | \$9 (5-13) |
| 50- 69 | 42,078 | \$147 (140-154) | 42,396 | \$119 (115-124) | \$27 (19-35) |
| 70 and above | 10,597 | \$574 (546-602) | 12,191 | \$377 (359-395) | \$197 (165-230) |
| **Terminal phase (60 days pre-death date)** |  |  |  |  |  |
| 18-29 | 33 | \$6,096 (3,123-9069) | <6 | \$1,051 (1450-3,554) | \$3,567.6 (-366-7,501) |
| 30-49 | 132 | \$10,951 (8,587-13,3145) | 111 | \$5809 (-26-11,645) | \$ 4,361 (-1,233-9,956) |
| 50- 69 | 834 | \$12,839 (12,021, 13,657) | 297 | \$5,213 (4, 591-5,835) | \$7,630 (6,594- 8,665) |
| 70 and above | 2,785 | \$7, 884 (7,575-8,193) | 3,480 | \$4, 231 (4, 087-4, 376) | \$3,657 (3,323- 3,991) |

n-sample size; CI-confidence interval; \*follow-up refers to one year after the index date or until death

Recent immigrants had lower costs in the pre-diagnosis, acute care, and post-acute care phases compared to long-term residents: pre-diagnosis $0 (-$9- $11) vs $11 ($6-$16), acute care $118 ($98- $137) vs $254 ($244-$263), and post-acute care phase $18 ($12-$24) vs $22 ($18-$26) (Table 5).

**Table 5:** Phase-specific COVID-19 per-person costs (2023 CAD) stratified by immigration status.

| **Immigration status** | **Exposed (n)** | **Exposed mean cost (95% CI)** | **Unexposed (n)** | **Unexposed mean cost (95% CI)** | **Attributable cost mean (95% CI)** |
| --- | --- | --- | --- | --- | --- |
| **Pre-diagnosis phase (20 days prior to index)** |  |  |  |  |  |
| Recent immigrant | 61,499 | \$87 (82-92) | 50,655 | \$75 (71-80) | \$12 (5-18) |
| Long term resident | 99,308 | \$126 (121-131) | 111,929 | \$113 (108-117) | \$13 (7-20) |
| **Acute phase (80 days post index)** |  |  |  |  |  |
| Recent immigrant | 61,447 | \$325 (310-339) | 50,648 | \$65 (62-68) | \$258 (243-273) |
| Long term resident | 99,177 | \$336 (325-348) | 111,881 | \$99 (96-103) | \$236 (225-248) |
| **Post acute (day 81 until end of follow up)** |  |  |  |  |  |
| Recent immigrant | 61,402 | \$97 (93-101) | 50,623 | \$67 (64-69) | \$30.3(25-35) |
| Long term resident | 98,949 | \$121 (117-125) | 111,710 | \$100 (98-103) | \$21 (16- 25) |

n-sample size; CI-confidence interval; \*follow-up refers to one year after the index date or until death (whichever is earlier)

Individuals living in the lowest-income neighbourhoods (quintile 1) had the highest mean cost in the pre-diagnosis ($135 [$126, $144]), acute ($405 [$384, $425]), and post-acute ($132 [$125, $138]) phases compared to other quintiles, while those living in the highest-income neighbourhoods (quintile 5) had the lowest costs (pre-diagnosis: $96 ($88-$105), acute: $285 ($263-$306), post-acute: $98 ($92-$105) (Table 6).

**Table 6:** Phase-specific COVID-19 per-person costs (2023 CAD) stratified by neighbourhood income quintile.

| **Neighbourhood income** | **Exposed (n)** | **Exposed mean cost (95% CI)** | **Unexposed (n)** | **Unexposed mean cost (95% CI)** | **Attributable mean cost (95% CI)** |
| --- | --- | --- | --- | --- | --- |
| **Pre-diagnosis phase (20 days prior to index)** |  |  |  |  |  |
| 1st quintile (lowest) | 39,207 | \$135 (126-144) | 39,707 | \$117 (110-123) | \$18 (7-29) |
| 2nd quintile | 34,637 | \$113 (105-120) | 35,048 | \$99 (92-106) | \$13 (3-24) |
| 3rd quintile | 34,652 | \$99 (93-106) | 35,045 | \$92 (85-99) | \$7 (-2-17) |
| 4th quintile | 28,470 | \$98 (90-107) | 28,712 | \$92 (85-99) | \$6 (-4-17) |
| 5th quintile | 22,834 | \$96 (88-105) | 23,055 | \$95 (86-105) | \$1(-11-14) |
| **Acute phase (80 days post index)** |  |  |  |  |  |
| 1st quintile (lowest) | 39,146 | \$405 (384-425) | 39,692 | \$108 (102-113) | \$297 (276-318) |
| 2nd quintile | 34,602 | \$343 (323-363) | 35,031 | \$89 (84-94) | \$254 (234-274) |
| 3rd quintile | 34,614 | \$302 (284-320) | 35,039 | \$85 (79-90) | \$217 (198-236) |
| 4th quintile | 28,449 | \$290 (270-309) | 28,708 | \$79 (74-84) | \$210 (190-230) |
| 5th quintile | 22,806 | \$285 (263-306) | 23,049 | \$82 (77-88) | \$202 (180-224) |
| **Post acute (day 81 until end of follow up)** |  |  |  |  |  |
| 1st quintile (lowest) | 39,076 | \$132 (125-138) | 39,632 | \$107 (102-112) | \$25 (17-33) |
| 2nd quintile | 34,534 | \$121 (113-129) | 34,978 | \$91 (87-95) | \$30 ((21-39) |
| 3rd quintile | 34,561 | \$101 (96-106) | 35,001 | \$87 (83-92) | \$14 (7-20) |
| 4th quintile | 28,414 | \$95 (89-100) | 28,672 | \$80 (76-85) | \$14 (7-21) |
| 5th quintile | 22,769 | \$98 (92-105) | 23,017 | \$83(78-87) | \$16 (8-24) |

n-sample size; CI-confidence interval; \*follow-up refers to one year after the index date or until death (whichever is earlier)

Individuals living in neighbourhoods with crowded housing (2 or more persons per room) had higher costs in the acute phase ($248 [$233-$263]) compared to those living in non-crowded housing neighbourhoods ($239 [$227- $251]) (Table 7)

**Table 7:** Phase-specific COVID-19 per-person costs (2023 CAD) stratified by household crowding index.

| Household crowding | Exposed (n) | Exposed mean cost (95% CI) | Unexposed (n) | Unexposed mean cost (95% CI) | Attributable mean cost (95% CI) |
| --- | --- | --- | --- | --- | --- |
| **Pre-diagnosis phase (20 days prior to index)** |  |  |  |  |  |
| One person or fewer per room | 91,441 | \$117 (112-122) | 92,479 | \$100 (96-104) | \$17 (10-23) |
| Two or more persons per room | 66,662 | \$101 (96-107) | 67,361 | \$98 (93-103) | \$3 (-3-11) |
| **Acute phase (80 days post index)** |  |  |  |  |  |
| One person or fewer per room | 91,347 | \$328 (317-340) | 92,445 | \$89 (86-92) | \$239 (227-251) |
| Two or more persons per room | 66,580 | \$335 (320-349) | 67,342 | \$86 (83-90) | \$248 (233-263) |
| **Post acute (day 81 until end of follow up)** |  |  |  |  |  |
| One person or fewer per room | 91,168 | \$114 (110-118) | 92,311 | \$91 (89-94) | \$22 (18-27) |
| Two or more persons per room | 66,496 | \$109 (104-114) | 67,270 | \$87 (84- 91) | \$21 (16-27) |

n-sample size; CI-confidence interval; \*follow-up refers to one year after the index date or until death (whichever is earlier)

No consistent directional trend was observed in phase-specific mean attributable costs by proportion of visible minorities or essential workers in a neighbourhood (Appendix Table A2 & A3)

The mean (minimum, maximum) total survival-adjusted 360-day cost per person was $3,297 ($3,096, $3,501), with males having higher costs ($3,618 [$3,316, $3,918]) than females ($2,727 [$2,478, $2,974]). Extrapolating to the population of 166,801 exposed individuals, the total survival-adjusted 360-day cost was estimated at $436 million ($408 –$462 million) (Table 8).

**Table 8.** Total COVID-19-attributable healthcare costs (2023 CAD), adjusted for survival, and stratified by sex.

|  | 30 days mean cost (min, max) | 90 days mean cost (min, max) | 180 days mean cost (min, max) | 360 days mean cost (min, max) |
| --- | --- | --- | --- | --- |
| **Total** | \$1,198 (1,155, 1,240) | \$ 2,148 (2,061, 2,234) | \$ 2,538 (2,412, 2,664) | \$ 3,297 (3,096, 3,501) |
| **Sex** |  |  |  |  |
| Female | \$ 933 (886, 971) | \$ 1,697 (1,599, 1,795) | \$ 2,038 (1,889, 2,185) | \$ 2,727 (2,478, 2,974) |
| Male | \$ 1,362 (1,299, 1,426) | \$ 2,435 (2,305, 2,565) | \$ 2,833 (2,645, 3,020) | \$3,618 (3,316, 3,918) |

## Discussion

Our study estimated healthcare costs attributable to confirmed COVID-19 in Ontario, Canada, over a 360-day period post-index, stratified by SDH.

Costs followed a typical U-shaped pattern (24): high during the acute phase, declining during the post-acute phase, and rising again during terminal care. Acute phase costs were the highest, consistent with the intensive resource demands of moderate to severe disease requiring hospitalization, ICU admission, and in some cases, mechanical ventilation (25).

Terminal phase costs for individuals with confirmed COVID-19 who died within 60 days of index (early deaths) were substantially higher than for those who died after 60 days (late deaths). Higher costs for early deaths likely reflect the use of resource-intensive interventions (e.g., mechanical ventilation and intensive care) during the acute phase of moderate to severe COVID-19 (26). In contrast, individuals with late deaths (>60 days) had longer hospital stays (median: 239 days [IQR: 55–627]), with hospitalization costs approximately 56.5% of those for early deaths, but incurred higher attributable home care and complex care costs, reflecting a shift to less costly care settings.

Compared to previous Ontario studies, our attributable mean cost estimate for the acute phase $244 was substantially lower than the $ 4,491 reported by Tsui *et al* (7) but similar to the data reported by Sander et al. $240 (9). The discrepancies between our findings and Tsui *et al*. could be due to differences in the costing window and the study time period. Tsui *et al.* estimated short-term COVID-19 healthcare costs using a 0–120-day costing window around the index date, based on cases from the first pandemic wave (January–June 2020). In contrast, both our study and Sander *et al*. used a much longer 360-day follow-up period with detailed 10-day costing intervals, capturing costs across multiple pandemic waves and enabling the inclusion of long-COVID and longer-term health-system impacts (7,9). Similar to Sander *et al*.(9), we observed sustained healthcare costs up to 360 days post-diagnosis, indicating a significant long-term burden of COVID-19. This aligns with evidence of persistent health issues associated with COVID-19, commonly referred to as post-COVID-19 condition, which lasts for more than 12 weeks after the initial infection (27). However, unlike Sander *et al.,* which focused on broader cost implications without stratifying by social determinants (9), our stratified analysis provides novel insights into how SDH were associated with variations in healthcare costs attributable to COVID-19. We found that sex, age, immigration status, income, and household crowding were associated with variation in phase-specific attributable costs. These findings align with Canadian epidemiological studies reporting higher rates of severe disease and mortality among these groups during the COVID-19 pandemic (13).

Multiple factors may explain the higher per-person healthcare costs observed among residents of low-income neighbourhoods. Individuals living in these areas often experience structural barriers to timely and effective healthcare, including longer travel times to healthcare facilities, fewer available healthcare providers, and limited access to preventive services (28). These barriers can potentially lead to delayed care with disease progressing to moderate or severe phase at the time of presentation, increasing reliance on acute-phase services such as emergency departments, hospitalizations, and ICU services (7,8). From our findings, hospitalization was the largest contributor to acute-phase costs among marginalized populations, including individuals living in low-income neighbourhoods. Consistent with our findings, a recent meta-analysis including over two million individuals reported higher odds of hospitalization among socioeconomically disadvantaged populations compared with advantaged groups during the COVID-19 pandemic (29).

Our study has some limitations. First, the C19INTGR dataset likely underestimates COVID-19 infections in Ontario due to incomplete testing, particularly during the early stages of the pandemic when testing was restricted to symptomatic individuals meeting specific criteria (30). As a result, the dataset potentially over-represents individuals with moderate to severe COVID-19 infections, who were more likely to be tested and incur higher healthcare costs, while those with asymptomatic or mild infections are underrepresented. Supporting this, a study conducted in Toronto, Ontario estimated that for each laboratory-confirmed symptomatic case between March and May 2020, there were approximately 18 undetected community infections (31), most of which were likely asymptomatic and required minimal to no healthcare services. Thus, our estimated attributable COVID-19 costs may be an overestimate. It is important to note that testing patterns varied by socioeconomic status: early in the pandemic, individuals in high-income neighbourhoods were more likely to be tested (4); however, testing later increased in low-income neighbourhoods due to higher infection rates and targeted public health efforts (4). This shift may have influenced our findings, as the initially higher testing rates in high-income neighbourhoods could have underestimated cost differences, while the subsequent increase in testing in low-income neighbourhoods could have contributed to greater healthcare use and costs observed in these populations. Second, our findings reflect the first pandemic year (2020) with limited preventive or treatment options and higher hospitalization rates, potentially overestimating costs compared to later waves characterized by vaccine and treatment availability, and milder variants. Furthermore, the area-level SDH variables (e.g., income, household crowding index, percentage of essential workers) were measured at the neighbourhood level using data at the DA level. While DA-level data serve as useful proxies in the absence of individual-level SDH data, they may not capture within-area and individual-level variability and could introduce bias. Therefore, our findings reflect area-level associations and should not be interpreted as individual-level effects. Lastly, while positive pre-diagnosis costs may reflect residual confounding bias, healthcare visits often preceded testing due to constrained testing capacity in 2020, aligning with early-pandemic healthcare-seeking patterns.

Our study has several strengths. We provide comprehensive estimates of the health and healthcare burden of confirmed COVID-19, stratified by key area-level SDH. Additionally, we matched exposed individuals to a historical unexposed cohort to account for healthcare delivery changes (e.g., surgery cancellations) in Ontario from May 2020 to February 2022, minimizing overestimation of COVID-19-attributable costs (14). We used a phase-of-care approach to mitigate the effect of censoring during the study follow-up period (32). Lastly, our phases of care periods and their associated costs can be mapped as health states in decision-analytic models to inform economic evaluations of COVID-19 interventions.

Our study highlights the substantial healthcare costs associated with COVID-19, particularly during acute and terminal phases, as well as the ongoing costs up to 360 days post-infection, indicating long-term sequelae. Our stratified analysis further supports the need for structural policy interventions that address the social and environmental conditions contributing to higher healthcare costs in marginalized communities. For example, enhancing healthcare infrastructure in low-income and crowded housing neighborhoods may reduce delays in care and prevent disease progression. Incorporating social risk indicators (e.g., income, housing density) into resource allocation frameworks can also help ensure that future health system planning and emergency responses are more equitable.

## Conclusion

Our study highlights significant healthcare costs attributable to confirmed COVID-19 in Ontario, concentrated in the acute and terminal phases and sustained up to 360 days post-infection. Costs varied by social determinants, with higher costs observed among marginalized populations, including those residing in low-income neighbourhoods, crowded housing, recent immigrants, and older adults. Our findings highlight systemic inequities in disease burden and resource utilization, emphasizing the need for early and tailored intervention strategies (e.g., vaccination, access to testing). Our findings can inform policies addressing existing disparities, optimizing resource distribution, and strengthening health system preparedness for future public health crises.

## Acknowledgements

This document used data adapted from the Statistics Canada 2016 Census Area Profiles and the Statistics Canada Postal CodeOM Conversion File, which is based on data licensed from Canada Post Corporation, and/or data adapted from the Ontario Ministry of Health Postal Code Conversion File, which contains data copied under license from ©Canada Post Corporation and Statistics Canada.

Parts of this material are based on data and/or information compiled and provided by Statistics Canada, the Canadian Institute for Health Information (CIHI), Ontario Health (OH), and the Ontario Ministry of Health, as well as Immigration, Refugees and Citizenship Canada (IRCC), current to September 2020. The analyses, conclusions, opinions, and statements expressed herein are solely those of the authors and do not reflect those of the funding or data sources, no endorsement is intended or should be inferred.

We thank the Toronto Community Health Profiles Partnership for providing access to the Ontario Marginalization Index. The authors thank Hong Lu for her coding contributions during the analysis and Patricia Aluko for proofreading the study manuscript.

## Author Contributions

AO and BS conceptualized the study. KL, SM, AO, and BS designed the study and methodology. AO and SS analyzed the data. AO drafted the manuscript. All authors reviewed, edited, and interpreted the data and approved the final manuscript. BS supervised the study.

## Data availability

The dataset from this study is held securely in a coded form at ICES. While legal data-sharing agreements between ICES and data providers (e.g., healthcare organizations and government) prohibit ICES from making the dataset publicly available, access may be granted to those who meet pre-specified criteria for confidential access, available at www.ices.on.ca/DAS. The full dataset creation plan and underlying analytic code are available from the authors upon request, with the understanding that computer programs may rely upon coding templates or macros that are unique to ICES and are therefore either inaccessible or may require modification.

## References

1. Public Health Ontario [Internet]. [cited 2024 Sept 5]. Ontario Respiratory Virus Tool. Available from: https://www.publichealthontario.ca/en/Data-and-Analysis/Infectious-Disease/Respiratory-Virus-Tool

2. Government of Ontario. 2021 Ontario Budget | Ontario.ca [Internet]. [cited 2021 Oct 19]. Available from: https://budget.ontario.ca/2021/index.html

3. https://www.fao-on.org. Financial Accountability Office of Ontario (FAO). [cited 2024 Sept 6]. Ontario Health Sector: 2023 Budget Spending Plan Review. Available from: https://www.fao-on.org/en/Blog/Publications/health-update-2023

4. Sundaram ME, Calzavara A, Mishra S, Kustra R, Chan AK, Hamilton MA, et al. Individual and social determinants of SARS-CoV-2 testing and positivity in Ontario, Canada: a population-wide study. CMAJ [Internet]. 2021 May 17 [cited 2024 Sept 12];193(20):E723–34. Available from: https://www.cmaj.ca/content/193/20/E723

5. Persaud N, Woods H, Workentin A, Adekoya I, Dunn JR, Hwang SW, et al. Recommendations for equitable COVID-19 pandemic recovery in Canada. CMAJ [Internet]. 2021 Dec 13 [cited 2022 Dec 28];193(49):E1878–88. Available from: https://www.cmaj.ca/content/193/49/E1878

6. Yang J, Andersen KM, Rai KK, Tritton T, Mugwagwa T, Reimbaeva M, et al. Healthcare resource utilisation and costs of hospitalisation and primary care among adults with COVID-19 in England: a population-based cohort study. BMJ Open. 2023 Dec 28;13(12):e075495.

7. Tsui TCO, Zeitouny S, Bremner KE, Cheung DC, Mulder C, Croxford R, et al. Initial health care costs for COVID-19 in British Columbia and Ontario, Canada: an interprovincial population-based cohort study. Canadian Medical Association Open Access Journal [Internet]. 2022 July 1 [cited 2024 Sept 6];10(3):E818–30. Available from: https://www.cmajopen.ca/content/10/3/E818

8. Sander B, Mishra S, Swayze S, Sahakyan Y, Duchen R, Quinn K, et al. Short-term and Long-Term Healthcare Costs Attributable to diagnosed COVID-19 in Ontario; Canada: A Population-Based Matched Cohort Study [Internet]. medRxiv; 2024 [cited 2024 Sept 12]. p. 2024.09.04.24313064. Available from: https://www.medrxiv.org/content/10.1101/2024.09.04.24313064v1

9. Sander B, Mishra S, Swayze S, Sahakyan Y, Duchen R, Quinn K, et al. Population-Based Matched Cohort Study of COVID-19 Healthcare Costs, Ontario, Canada. Emerg Infect Dis [Internet]. 2025 Apr [cited 2025 Aug 5];31(4):710–9. Available from: https://www.ncbi.nlm.nih.gov/pmc/articles/PMC11950279/

10. RECORD Reporting Guidelines [Internet]. [cited 2024 Sept 18]. Available from: https://www.record-statement.org/

11. ICES. ICES Data [Internet]. [cited 2021 Oct 21]. Available from: https://www.ices.on.ca/Data-and-Privacy/ICES-data

12. ICES [Internet]. [cited 2024 Sept 18]. ICES. Available from: https://www.ices.on.ca/

13. Public Health Agency of Canada. From risk to resilience: An equity approach to COVID-19 – The Chief Public Health Officer of Canada’s Report on the State of Public Health in Canada 2020 [Internet]. 2020 [cited 2021 June 17]. Available from: https://www.canada.ca/en/public-health/corporate/publications/chief-public-health-officer-reports-state-public-health-canada/from-risk-resilience-equity-approach-covid-19.html

14. Crawley M. Ontario orders hospitals to halt non-emergency surgeries as COVID-19 patients fill ICUs. CBC News [Internet]. 2021 Apr 9 [cited 2024 Nov 23]; Available from: https://www.cbc.ca/news/canada/toronto/covid-19-ontario-hospitals-elective-surgery-icu-patients-1.5980755

15. ICES Data Dictionary [Internet]. [cited 2024 Nov 23]. Available from: https://datadictionary.ices.on.ca/Applications/DataDictionary/Default.aspx

16. The Johns Hopkins ACG® System, Technical Reference Guide, Version 10.0, December 2011. :264.

17. Sundaram M, Calzavara A, Mishra S, Kustra R, Chan A, Hamilton M, et al. The Individual and Social Determinants of COVID-19 Diagnosis in Ontario, Canada: A Population-Wide Stud [Internet]. 2020 [cited 2024 Sept 12]. Available from: https://europepmc.org/article/PPR/PPR237585

18. Canada PHA of. Summary: The Direct Economic Burden of Socio-Economic Health Inequalities in Canada [Internet]. 2016 [cited 2024 Nov 23]. Available from: https://www.canada.ca/en/public-health/services/publications/science-research-data/summary-direct-economic-burden-socio-economic-health-inequalities-canada.html

19. Fitzpatrick T, Rosella LC, Calzavara A, Petch J, Pinto AD, Manson H, et al. Looking Beyond Income and Education: Socioeconomic Status Gradients Among Future High-Cost Users of Health Care. American Journal of Preventive Medicine [Internet]. 2015 Aug 1 [cited 2025 July 2];49(2):161–71. Available from: https://www.sciencedirect.com/science/article/pii/S0749379715000823

20. Government of Canada SC. 2016 Census Program [Internet]. 2016 [cited 2024 Dec 28]. Available from: https://www12.statcan.gc.ca/census-recensement/2016/ref/dict/geo021-eng.cfm

21. Austin PC. Balance diagnostics for comparing the distribution of baseline covariates between treatment groups in propensity-score matched samples. Stat Med [Internet]. 2009 Nov 10 [cited 2021 Apr 11];28(25):3083–107. Available from: https://www.ncbi.nlm.nih.gov/pmc/articles/PMC3472075/

22. Post COVID-19 condition (Long COVID) [Internet]. [cited 2024 Sept 20]. Available from: https://www.who.int/europe/news-room/fact-sheets/item/post-covid-19-condition

23. Yabroff KR, Lamont EB, Mariotto A, Warren JL, Topor M, Meekins A, et al. Cost of care for elderly cancer patients in the United States. J Natl Cancer Inst. 2008 May 7;100(9):630–41.

24. Shing E, Wang J, Nelder MP, Parpia C, Gubbay JB, Loeb M, et al. The direct healthcare costs attributable to West Nile virus illness in Ontario, Canada: a population-based cohort study using laboratory and health administrative data. BMC Infect Dis [Internet]. 2019 Dec 17 [cited 2024 Dec 2];19:1059. Available from: https://www.ncbi.nlm.nih.gov/pmc/articles/PMC6918579/

25. Kaier K, Heister T, Motschall E, Hehn P, Bluhmki T, Wolkewitz M. Impact of mechanical ventilation on the daily costs of ICU care: a systematic review and meta regression. Epidemiol Infect [Internet]. [cited 2024 Dec 2];147:e314. Available from: https://www.ncbi.nlm.nih.gov/pmc/articles/PMC7003623/

26. Mac S, Barrett K, Khan YA, Naimark DM, Rosella L, Ximenes R, et al. COVID-19 Demographics, Acute Care Resource Use and Mortality by Age and Sex in Ontario, Canada: Population-based Retrospective Cohort Analysis [Internet]. Infectious Diseases (except HIV/AIDS); 2020 Nov [cited 2021 Mar 16]. Available from: http://medrxiv.org/lookup/doi/10.1101/2020.11.04.20225474

27. Canada PHA of. Post-COVID-19 condition (long COVID) [Internet]. 2021 [cited 2024 Dec 5]. Available from: https://www.canada.ca/en/public-health/services/diseases/2019-novel-coronavirus-infection/symptoms/post-covid-19-condition.html

28. Coombs NC, Campbell DG, Caringi J. A qualitative study of rural healthcare providers’ views of social, cultural, and programmatic barriers to healthcare access. BMC Health Services Research [Internet]. 2022 Apr 2 [cited 2025 Nov 22];22(1):438. Available from: 10.1186/s12913-022-07829-2

29. Zhu YJ, Tang K, Zhao FJ, Yu BY, Liu TT, Zhang LL. Impact of Social Deprivation on Hospitalization and Intensive Care Unit Admission among COVID-19 Patients: A Systematic Review and Meta-Analysis. Iran J Public Health. 2022 Nov;51(11):2458–71.

30. Item-4-Provincial-Testing-Guidance-Update.pdf [Internet]. [cited 2025 Aug 5]. Available from: https://www.corhealthontario.ca/Item-4-Provincial-Testing-Guidance-Update.pdf

31. Desta BN, Ota S, Gournis E, Pires SM, Greer AL, Dodd W, et al. Estimating the Under-ascertainment of COVID-19 cases in Toronto, Ontario, March to May 2020. J Public Health Res [Internet]. 2023 May 12 [cited 2025 Aug 5];12(2):22799036231174133. Available from: https://www.ncbi.nlm.nih.gov/pmc/articles/PMC10184215/

32. Wijeysundera HC, Wang X, Tomlinson G, Ko DT, Krahn MD. Techniques for estimating health care costs with censored data: an overview for the health services researcher. Clinicoecon Outcomes Res [Internet]. 2012 June 1 [cited 2024 Sept 20];4:145–55. Available from: https://www.ncbi.nlm.nih.gov/pmc/articles/PMC3377439/

